# African ancestry-enriched variants in the *GATM* gene are associated with elevated serum creatinine levels

**DOI:** 10.1101/2025.03.07.25323581

**Authors:** Shivam Sharma, Courtney A. Astore, Leonardo Mariño-Ramírez, I. King Jordan

## Abstract

**Background:** Serum creatinine (Scr) levels are routinely used to estimate kidney function and health. Individuals of African ancestry have higher Scr levels – controlling for differences in age, sex, size, kidney function, and disease status – compared to individuals from other ancestral backgrounds. The reason for this difference is unknown. We hypothesized that there may be genetic variants found at relatively high frequency in African ancestry groups (African ancestry-enriched variants) that are associated with elevated Scr levels African ancestry individuals.

**Methods:** Our study sample is made up of participants from the All of Us Research Program. We used whole genome sequence data to estimate genetic ancestry for All of Us participants and selected a cohort of 18,979 participants with two way African-European admixture, available Scr level measures, and demographic covariables. We performed a series of ancestry-informed association studies of Scr levels on this cohort to test our hypothesis of African ancestry-enriched variants associated with Scr.

**Results:** Study participants show an average of 80.8% African and 17.5% European ancestry. Participant Scr levels are positively correlated with African ancestry for females (*ρ*=0.79) and males (*ρ*=0.84). The same peak of genome-wide significant associations was identified on chromosome 15 (15q23:45.3Mb-45.5Mb) using standard GWAS, haplotype-based admixture mapping, and ancestry-specific GWAS. The alternate allele for the lead GWAS variant (rs2467850, chr15:45379909:C:T) is positively associated with Scr levels (*β*=0.07, *p*=2.28×10^−17^) and found at higher frequency in African (0.413) compared to European ancestry (0.001) groups. Fine mapping identified a credible set of 14 variants co-located with the *GATM* gene, which encodes a biosynthetic enzyme for creatine, a metabolic precursor of creatinine. 13 of these variants are positively associated with *GATM* expression, based on a previous study of whole blood eQTL in African Americans, and they all show similar patterns of African ancestry-enrichment. An Scr polygenic score based on 10 African ancestry-enriched variants completely attenuates the observed association of African ancestry with Scr levels.

**Conclusions:** Our findings indicate that African ancestry-enriched variants up-regulate the *GATM*, thereby explaining the higher levels of Scr observed in individuals of African ancestry, and underscore the potential for using genetic data to better calibrate kidney function equations.

## Background

Creatine synthesis begins in the liver, kidneys, or pancreas, and the synthesized creatine is subsequently transported to skeletal and heart muscle, where it is phosphorylated to serve as an energy source [1]. Creatine is catabolized in muscle tissue to produce creatinine, a waste product that gets filtered out of the blood by the kidneys [2]. Serum creatinine (Scr) levels are used together with demographic variables to assess kidney function by calculating the estimated glomerular filtration rate (eGFR). Scr levels have been found to vary with age, sex, and race; African American patients have higher average Scr levels, at the same measured glomerular filtration rate (mGFR), compared to patients from other groups [3, 4]. Accordingly, eGFR equations, such as the 1999 Modification of Diet in Renal Disease (MDRD) and 2009 Chronic Kidney Disease Epidemiology Collaboration (CKD-EPI) equations, previously included age, sex, and race coefficients along with patient Scr levels [5, 6]. Inclusion of the race coefficient in the CKD-EPI equation was shown to increase the accuracy of the eGFR equation for African American patients [7, 8]. However, the inclusion of race in eGFR equations was widely debated, since race is considered to be a social rather than a biological category [9, 10]. Eventually, in 2021, the National Kidney Foundation– American Society of Nephrology Task Force recommended the removal of race from eGFR calculations, and the currently used CKD-EPI Scr eGFR equation does not include a race coefficient [11, 12].

One of the concerns about the inclusion of patient race in the previous eGFR equations was the lack of any known mechanism that could explain why considering race improved the eGFR equation performance [8]. It has long been assumed that differences in Scr levels are influenced by variations in muscle mass and diet; however, the inclusion of height and weight in the CKD-EPI equation did not attenuate the effect of race or meaningfully improve its performance [7]. In addition, race is socially defined and thus assumed to be a poor proxy for patient biological differences [13]. Genetic ancestry, on the other hand, is a characteristic of the genome and manifests as allele frequency differences between population groups [14]. In our previous study of the UK Biobank, we found that percentage of African genetic ancestry is strongly and positively correlated with Scr levels, irrespective of other demographic and socioeconomic variables [4]. In the US, replacing patient race with African genetic ancestry in the CKD-EPI equation resulted in similar eGFR estimates [15].

We hypothesized that there may be African ancestry-enriched genetic variants or haplotypes that are associated with elevated Scr levels in individuals of African ancestry, and we performed a series of ancestry-informed association studies of Scr levels to test this hypothesis. Our approach relies on the fact that individuals with African ancestry in the US tend to be admixed with ∼80% African and 20% European genetic ancestry on average, which allowed us to stratify variants by ancestry on a shared genetic background. The study cohort is made up of participants from the All of Us Research Program (All of Us), a large-scale biomedical dataset that aims to reflect the diversity of US population, who show a continuum of African-European admixture and do not have impaired kidney function [16]. We used a combination of ancestry-stratified and local-ancestry enabled GWAS, together with admixture mapping, to screen for ancestry-specific Scr genetic associations in the All of Us study cohort.

## Methods

### Study cohort and data

This study was performed on the All of Us controlled tier dataset version 7 (curated version C2022Q4R9). Participant inclusion criteria and analysis steps are shown in Fig. S1. Continental genetic ancestry percentages were inferred for 297,549 unrelated All of Us participants using participant Illumina Global Diversity array data merged with variant data from global references populations and the program Rye [17]. Detailed steps for reference panel selection, variant quality control, merging, and harmonization, followed by genetic ancestry inference are described in our previous work [18]. A total of 60,132 participants with two-way admixed African-European ancestry were selected using the criteria: African > 5% and European < 95%, with the combined contribution from other ancestry groups restricted to a maximum of 5%.

Scr values and chronic kidney disease status were obtained for participants through their electronic health records (EHR). Since this study focused on healthy individuals, participants with chronic kidney disease (any stage) or hypertensive chronic kidney disease were removed using the phecodes 585 and 401.22 respectively [19]. Scr values were extracted for the remaining 27,086 participants and harmonized to *mg/dL* units where possible. Scr interquartile range was defined within and across participant EHR to remove outlier measurements to yield a final cohort of 24,272 participants. Participant Scr levels were inverse normal transformed for use in all association analyses.

### Local genetic ancestry

Local ancestry tracts were inferred for 21,926 two-way African-European admixed participants who had serum creatinine levels and array data. The ancestry reference panel was created using 372 European (CEU, GBR, IBS, and TSI) and 397 African (ESN, GWD, MSL, and YRI) genome samples from the 1000 Genomes project [20]. A total of 947,835 variants were extracted from the merged array data and further filtered to retain variants with a minor allele frequency of at least 0.1% and a minimum count of 50. This dataset was phased using Beagle 5.4 with no imputation and the number of iterations increased to 15 [21]. African and European local ancestry tracts were subsequently inferred using Gnomix with *smooth_size* reduced to 45 and up to 20 generations were simulated for the training model [22].

### Genome-wide association testing (GWAS)

We used short-read whole genome sequencing (srWGS) variant data from the Allele Count/Allele Frequency (ACAF) threshold callset for genome-wide single variant association testing. For the remaining 18,979 participants who had srWGS-ACAF data, we applied additional variant-level filtering using Plink v2 to exclude variants with a minor allele frequency below 1%, a Hardy-Weinberg equilibrium deviation p-value less than 10^−15^, or a missingness rate exceeding 10% to yield a total of 17,689,489 variants [23]. Variant-Scr association testing was performed using a linear mixed model as implemented in SAIGE v1.3.1 for single variant testing [24]. Age, sex, age^2^, and the first ten principal components were used as covariates in the model to correct for fixed effects, while random effects were accounted for using a genomic relationship matrix to control for cryptic relatedness among participants. As suggested by SAIGE authors, variants with minor allele counts less than 20 were removed from the analysis. LD-based clumping was performed using Plink v2 followed by conditional analysis through SAIGE, executed for chromosome 15, using the *condi4on* flag where LD-independent SNPs were fed into the testing model.

### Admixture mapping

Local ancestry haplotype dosages for African ancestry were used to perform a genome-wide admixture mapping scan, which identified genomic regions where African ancestry is associated with Scr levels. Admixture mapping was performed using the GENESIS v2.30.0, with a GDS data frame prepared using the gdsfmt v1.42.0 [25, 26]. The analysis involved fitting a null model using the *fitNullModel* to account for covariates (age, sex, principal components one to ten), followed by applying the *admixMap* function to identify associations between local ancestry and Scr levels. The genome-wide significance threshold was calculated using STEAM v0.1.0, where number of generations since admixture were inferred as 8, and threshold was determined as 3×10^−5^ [27]. An additional conditional admixture mapping was carried out on srWGS inferred local ancestry from chromosome 15 with lead SNP added to the list of covariates.

### Ancestry-specific GWAS

Local-ancestry boosted GWAS, was performed using Tractor on chromosome 15 to explore ancestry-specific genetic contributions to Scr levels [28]. Local ancestry was inferred from srWGS data for 725 ancestral haplotypes encompassing 459,269 variants. The analysis incorporated covariates including age, age^2^, sex, and the first ten principal components to account for confounding effects.

### Variant fine mapping and polygenic scores

Linkage disequilibrium (LD) correlation analysis was performed using Plink v2 to identify patterns of LD 250kb up or down-stream of the lead variant [23]. Fine mapping of associated region was conducted using susieR, a Bayesian regression framework that estimated credible sets of variants, 250kb up or down-stream of the lead SNP, likely to contain causal signals [29]. Locus zoom and variant manhattan plots were made using R packages dplyr, ggplot2, and ggrepel. Polygenic scores were calculated for participants using PRSice-2 with default settings for quantitative traits [30].

## Results

### Variation of Scr levels with African ancestry

The study cohort is made up of 18,979 All of Us participants, who are 65.1% female and 34.9% male, with an average age of 54 years. Participants show a continuum of genetic diversity, falling between African and European reference populations along principal component 1 (PC1) and dispersed along PC2 (Fig. 1A). Participant genetic ancestry percentages are also continuously distributed, with an average of 80.8% African and 17.5% European ancestry (Fig. 1B). Scr levels were approximately normally distributed for both males and females, with males showing higher overall Scr levels, consistent with previous findings (Fig. S2) [4, 31, 32]. Scr levels are positively correlated with African ancestry, *ρ* = 0.79 for females and *ρ* = 0.84 for males, consistent with previous studies (Fig. 1C) [4, 15]. Adjusting for sex and age, African ancestry is significantly associated with Scr levels (*β*=0.09, *p*=1.65×10^−34^; Fig. 1D).

**Figure 1.**
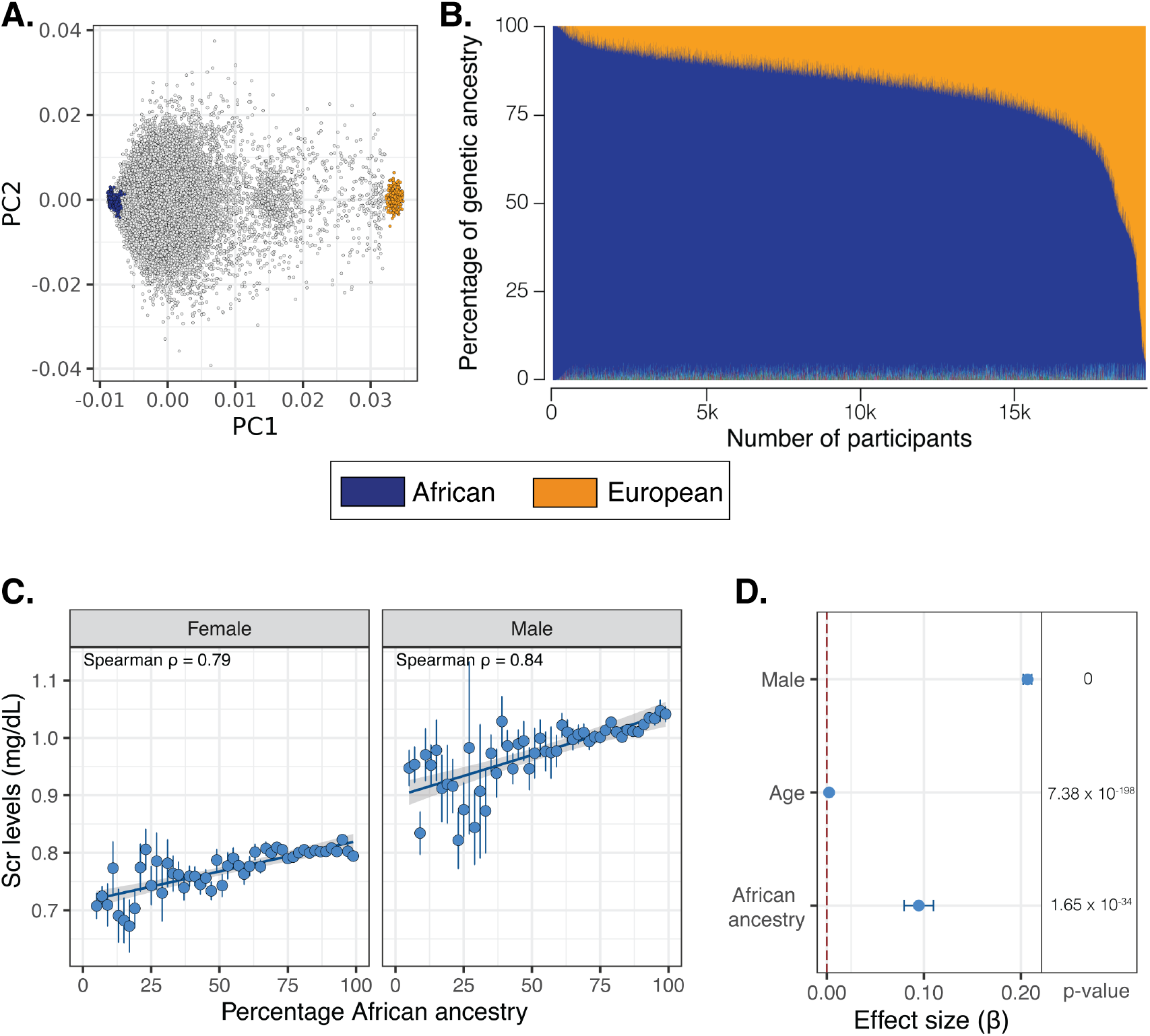
Genetic ancestry and Scr levels. (A) Genomic PCA for All of Us participants (grey) with African (blue) and European (orange) reference populations. (B) Participant African (blue) and European (orange) ancestry percentages. (D) Correlation of Scr levels and African ancestry stratified by sex. Average±SE Scr levels are shown for 50 ancestry percentage bins. Linear regression line is shown with 95% CI. (D) Scr association effect sizes ±95% CI, along with p-values, show for sex, age, and African ancestry.

### Scr GWAS

A GWAS of Scr levels was conducted using SAIGE for single-variant association tests of 17.6 million variants (referred to hereafter as ‘standard GWAS’ to distinguish from other methods). The GWAS Q-Q plot and *λ*_GC_ = 1.04 do not show evidence of genomic inflation owing to population structure (Fig. S3). A major genome-wide significant peak of variants was identified on chromosome 15 (15q23:45.3Mb-45.5Mb), with a secondary, less significant peak on chromosome 11 (11p15.4:30.7Mb) (Fig. 2A). LD-based clumping of these peaks identified three independent loci, two on chromosome 15 and one on chromosome 11 (Table 1).

**Table 1.** Scr-associated loci. Lead variant, or haplotype, summary statistics are shown for standard GWAS, admixture mapping, and ancestry-specific GWAS.

| Standard GWAS |  |  |  |  |  |  |  |  |  |  |
| --- | --- | --- | --- | --- | --- | --- | --- | --- | --- | --- |
| ID | CHR | POS | REF | ALT | ALT<sub>Freq</sub> | BETA | SE | p | AFR<sub>Freq</sub> | EUR<sub>Freq</sub> |
| rs2467850 | 15 | 45379909 | C | T | 0.414 | 0.074 | 0.0087 | 2.28×10<sup>-17</sup> | 0.413 | 0.001 |
| rs34830202 | 11 | 30755467 | G | GA | 0.127 | -0.074 | 0.0131 | 1.73×10<sup>-8</sup> | 0.125 | 0.461 |
| rs55673230 | 15 | 45466280 | A | G | 0.126 | -0.074 | 0.0131 | 1.80 ×10<sup>-8</sup> | 0.122 | 0.475 |
| Admixture mapping |  |  |  |  |  |  |  |  |  |  |
| CHR | POS |  | BETA |  | SE | p |  |  |  |  |
| 15 | 45.3M - 45.4M |  | 0.056 |  | 0.0109 | 2.57×10<sup>-07</sup> |  |  |  |  |
| Ancestry-specific GWAS |  |  |  |  |  |  |  |  |  |  |
| ID | CHR | POS | REF | ALT | ALT<sub>Freq</sub> | BETA | SE | p | AFR<sub>Freq</sub> | EUR<sub>Freq</sub> |
| rs1153848 | 15 | 45403676 | A | G | 0.512 | 0.069 | 0.0094 | 2.28×10<sup>-13</sup> | 0.414 | 0.001 |

**Figure 2.**
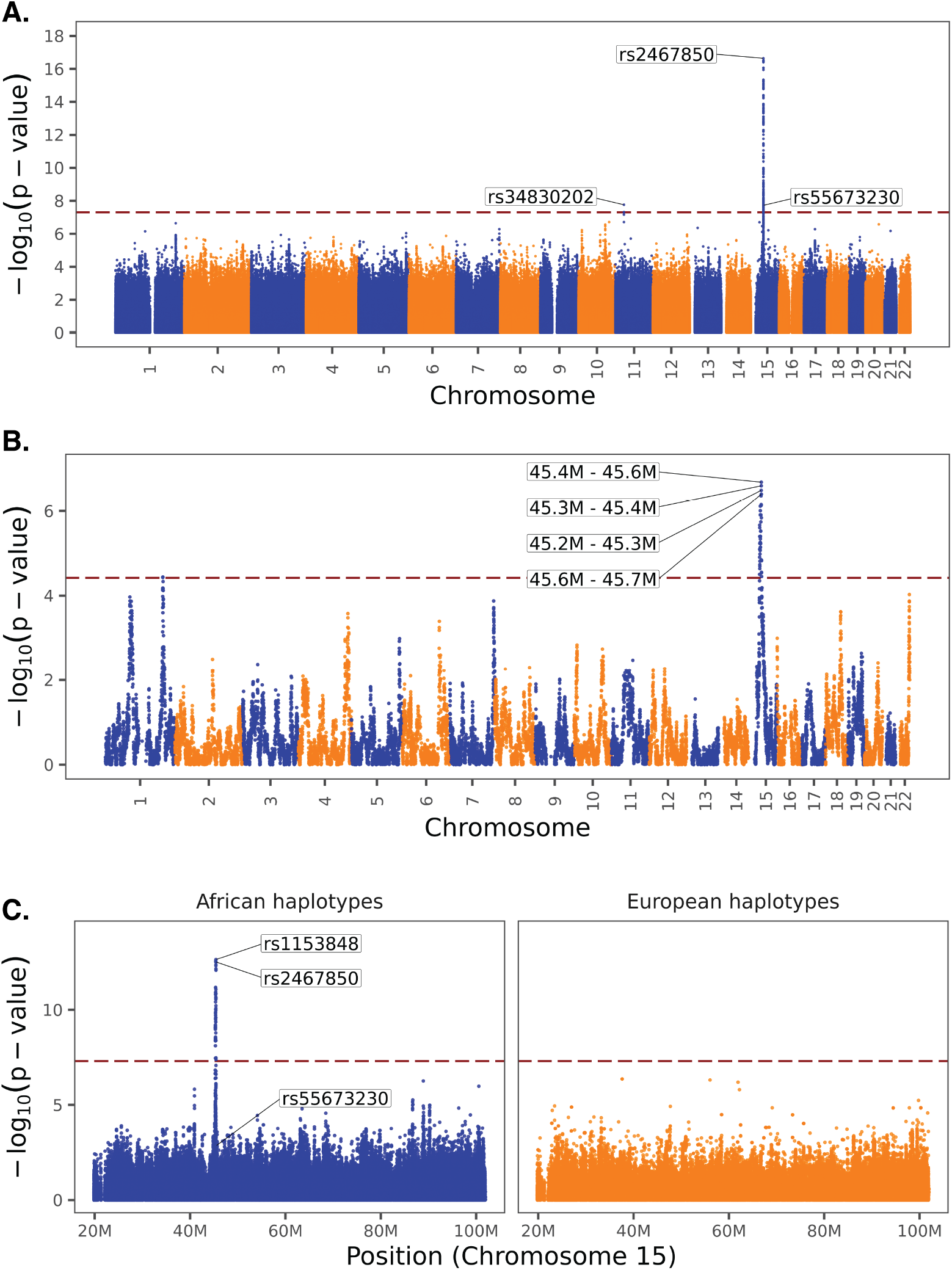
Genome-wide association testing for Scr levels. Manhattan plots showing the significance (−log10 p-values) of Scr-variant (haplotype) associations are shown for (A) standard GWAS, (B) admixture mapping, and (C) ancestry-specific GWAS.

The lead variant on chromosome 15 (rs2467850) is positively associated with Scr levels (*β*=0.07, *p*=2.28×10^−17^) and shows an alternate allele frequency of 0.41 in the study cohort (*ALT*_*Freq*_) and 0.41 in the Gnomad African/African American genetic ancestry group (*AFR*_*Freq*_), compared to an allele frequency of 0.001 in the Gnomad European (non-Finnish) group (*EUR*_*Freq*_). The second variant on chromosome 15 (rs55673230) is negatively associated with Scr levels (*β*=-0.07, *p*=1.80×10^−8^), with *ALT*_*Freq*_=0.13, *AFR*_*Freq*_=0.13, and *EUR*_*Freq*_=0.46. The lead variant on chromosome 11 (rs34830202) is negatively associated with Scr levels (*β*=-0.07, *p*=1.73×10^−8^), with *ALT*_*Freq*_=0.13, *AFR*_*Freq*_=0.12, and *EUR*_*Freq*_=0.48. The associated loci on chromosome 15 loses significance only when conditioned on lead variant rs2467850, and with second independent variant rs55673230 (Fig. S4).

In summary, all three of these lead variants show large allele frequency differences between African and European ancestry groups, with the lead variant on chromosome 15 being essentially monomorphic for the reference allele in the European ancestry group. Given the ancestry-specific allele frequencies for these variants, and the direction of their effects, they all contribute to African ancestry associated increases in Scr levels in the All of Us study cohort.

### Scr admixture mapping

A genome-wide admixture mapping scan was conducted using GENESIS to identify ancestry-specific associations between Scr levels and African ancestry haplotypes. A total of 17,693 haplotypes were tested, and a genome-wide significant African ancestry association was found the same chromosome 15 locus discovered by GWAS (15q23:45.3Mb-45.5Mb; Fig. 2B). This result supports the association of African ancestry enriched variants with increased Scr levels among individuals with African-European admixture. Conditional admixture mapping analysis was performed on chromosome 15, including the GWAS lead variant rs2467850 as an additional covariate. This adjustment resulted in the loss of significance for the admixture mapping signal, suggesting that the association in this region is largely driven by rs2467850 or variants in strong linkage disequilibrium with it (Fig. S5).

### Scr ancestry-specific GWAS

An ancestry-specific GWAS of Scr levels was performed for chromosome 15 (20Mb-100Mb) using Tractor. Tractor integrates variant data with local ancestry assignments of African and European ancestry haplotypes to calculate ancestry-specific associations. A single genome-wide significant peak including four African ancestry-specific variant associations with Scr levels was identified spanning the same chromosome 15 region (15q23:45.2Mb-45.7Mb) discovered by standard GWAS and admixture mapping (Fig. 2C). There were no significant European ancestry-specific associations seen for chromosome 15. The lead African ancestry-specific variant association on chromosome 15 (rs55673230; *β*=0.07, *p*=2.28×10^−13^) is in linkage disequilibrium with the lead variant discovered by standard GWAS (rs2467850), which was the third most significant African ancestry-specific variant association in this region (*β*=0.07, *p*=3.01×10^−13^). The second most significant variant association in the standard GWAS (rs55673230), which is unlinked to the lead variant and showed a negative effect size, did not reach genome-wide significance in either the African or European ancestry-specific GWAS.

### Chromosome 15 variant fine-mapping

Fine mapping of Scr associated variants on chromosome 15 was conducted using susieR for both the standard and ancestry-specific GWAS results. Fine mapping for the standard GWAS results identified a single credible set of 14 variants, which is likely (95% posterior inclusion probability) to contain one or more causal variants responsible for the observed association with Scr (Fig. 3A). Both the lead variant from the standard GWAS (rs2467850) and the linked top variant from the African ancestry-specific GWAS (rs1153848) are present within the credible set. The second, unlinked lead variant from standard GWAS (rs55673230) was absent from the credible set. Fine mapping of the African ancestry-specific GWAS results identified a single credible set of 12 variants, which are a subset of the GWAS credible set and include both rs2467850 and rs55673230 (Fig. 3B). The credible variant set is centered on the *GATM* gene, which encodes a creatinine biosynthesis enzyme, with the standard GWAS lead variant (rs2467850) is located 1,365bp upstream of the *GATM* transcription start site.

**Figure 3.**
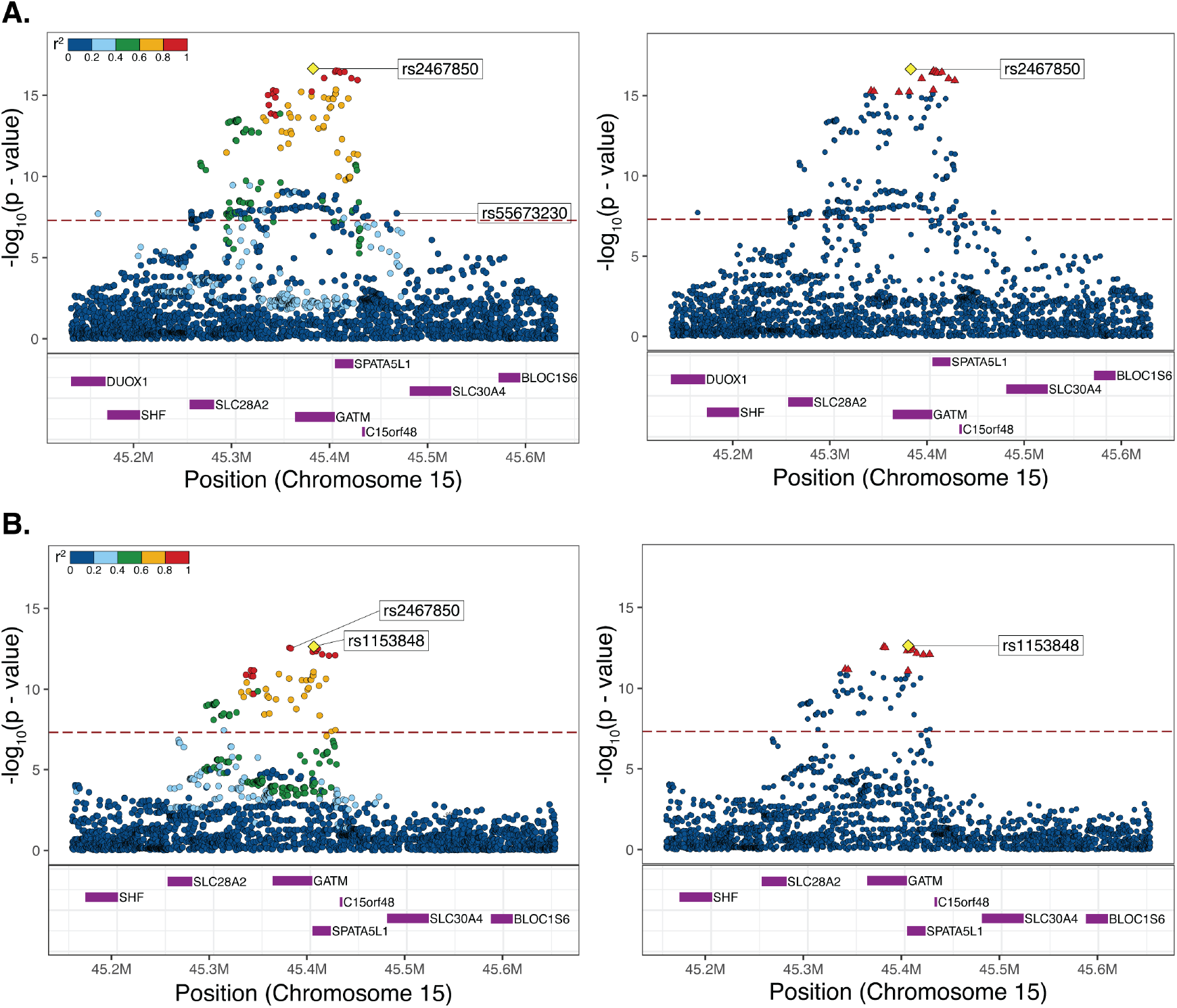
Variant fine mapping. Regional association plots showing LD correlation patterns (left) and fine-mapping (right) for chromosome 15q23. Lead variants are shown as yellow diamonds. Variants are color-coded by LD (r^2^) in the left panel, and credible variants are shown as red triangles in the right panel. Results are shown for (A) standard GWAS and (B) African ancestry-specific GWAS.

Inspection of cis eQTL characterized from whole blood in 757 African Americans revealed that 13 of the 14 *GATM* credible variants, with one missing from the eQTL data set, have alternate alleles that are positively associated with *GATM* expression (Table S1) [33]. All of these *GATM* expression associated alleles are African ancestry-enriched. Considered together, the GWAS fine mapping and eQTL data indicate that the African ancestry-enriched Scr associated variants discovered here exert a causal effect on up-regulation of the creatine biosynthesis *GATM* gene, thereby leading to the higher observed levels of Scr in participants with African ancestry.

### African ancestry-enriched variants explain Scr association with African ancestry

The observed association of African ancestry with Scr levels was conditioned on Scr GWAS discovered variants. To do so, a series of Scr polygenic scores (*PGS*_*Scr*_) were generated from the standard GWAS results by including variants that are both African ancestry-enriched and Scr-associated: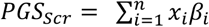, where *n* = 1 − 14 variants, *x*_*i*_ is the alternate allele dosage for variant *i*, and *β*_*i*_ is the effect size for variant *i*. The lead variants from unlinked GWAS loci were used to create the *PGS*_*Scr*_, and African ancestry-enrichment was quantified by taking the difference between African and European allele frequencies: Δ*Allele*_*Freq*_ = *AFR*_*Freq*_ − *EUR*_*Freq*_. Variants were prioritized and sorted in descending order, based on the product of their effect size (*β*), significance level (−*log*_10_*p*), and Δ*Allele*_*Freq*_. The number of variants (*n*) needed to attenuate the observed correlation between Scr levels and African ancestry was evaluated by comparing estimated African ancestry effect sizes (*β*_*AFR*_) for adjusted models of Scr levels: *Scr*∼*AFR* + *PRS*_*Scr*_ + *age* + *sex*. With zero variants included in the *PRS*_*Scr*_, the Scr African ancestry effect size *β*_*AFR*_=0.09 (as in Fig. 1D). Estimated *β*_*AFR*_ values decrease with increasing numbers of variants included in the *PRS*_*Scr*_ (Fig. 4A). The *β*_*AFR*_ becomes non-significant when 10 variants included in *PGS*_*Scr*_ (*β*_*AFR*_=0.01, *p*=0.17; Fig. 4B).

**Figure 4.**
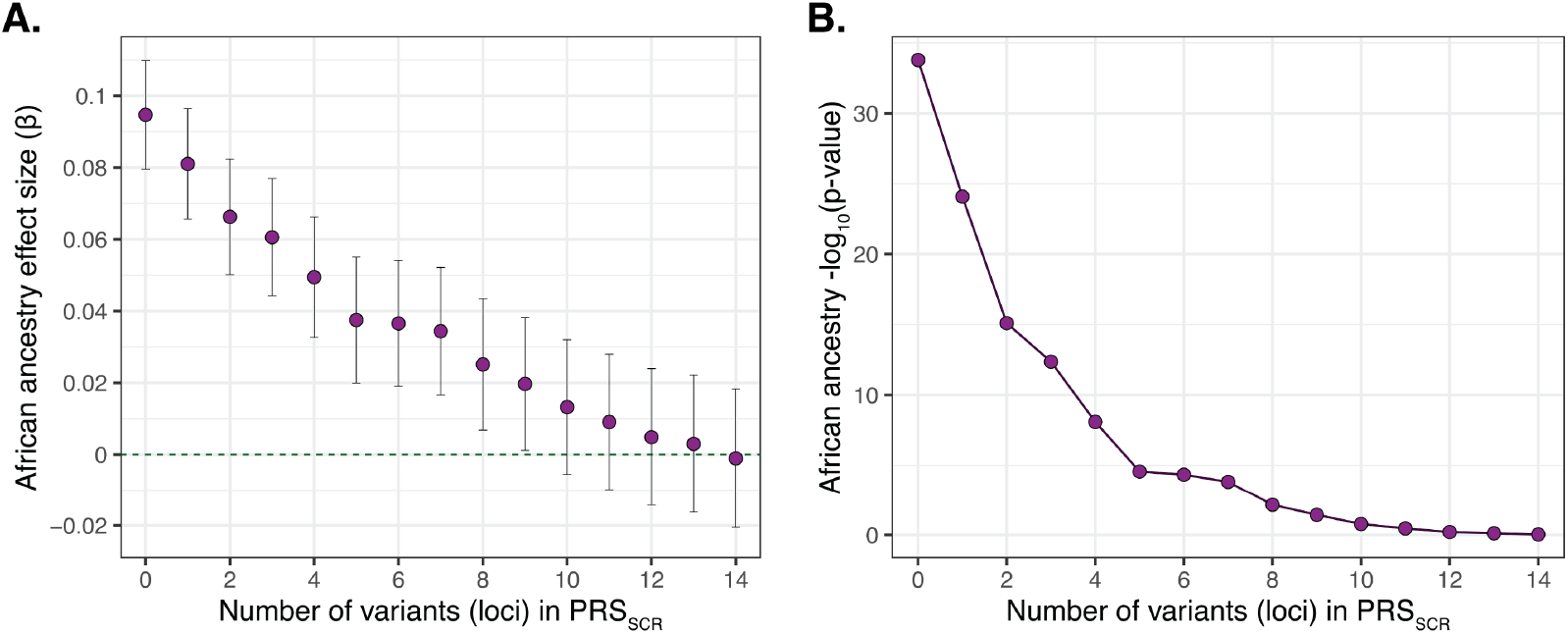
Conditional Scr African ancestry association. African ancestry association is attenuated with each additional variant included in the Scr polygenic score (PRSSCR). (A) Scr African ancestry effect sizes (*β*) compared to the number of variants in included in PRSSCR. (B) Significance of the Scr African ancestry association compared to the number of variants in included in the PRSSCR.

## Discussion

Ancestry-linked differences in Scr levels have been at the heart of a vigorous debate on the use of race in clinical algorithms [34]. Race was previously included in eGFR equations, owing to greater prediction accuracy in African ancestry patients [5, 6]. The use of race in eGFR equations was vigorously contested, primarily on racial justice grounds, and eventually overturned in 2021 [8, 12]. A key question that was never resolved during this debate, was the reason for higher observed Scr levels among African ancestry patients. To date, there is no known variable or mechanism that explains the observed correlation between African ancestry and Scr levels.

Here, using a series of ancestry-stratified association studies, we were able to discover a genetic basis for the higher levels of Scr among African ancestry patients. We found a highly significant association between Scr levels and African ancestry-enriched variants at chromosome 15 (15q23:45.3Mb-45.5Mb), which replicates across three different association methods (Figure 2). The lead GWAS variant is found at >40% frequency in African ancestry groups and is virtually absent in European ancestry groups (Table 1). The correlation between Scr and African ancestry can be completely attenuated by a polygenic score consisting of just 10 variants (PGSSCR; Figure 4). The variant peak, and the smaller credible set of variants, are centered on the *GATM* gene, with the lead variant located in the proximal promoter region upstream of the transcription start site (Figure 3). The putatively causal variants in the credible set are all African ancestry-enriched and positively associated with *GATM* expression levels (Table S1). *GATM* encodes glycine amidinotransferase, a key enzyme in the creatine biosynthesis pathway. Creatinine is a waste product of creatine catabolism, which is filtered out of the blood by the kidneys, thus linking *GATM* directly to Scr levels.

According to the GWAS catalog, 14 previous studies have reported Scr or eGFR variant associations at or near the *GATM* locus, and 8 of those studies included African ancestry participants [35-48]. We compared the results of those previous studies with our own findings, focusing on ancestry-specific allele frequencies (*AFR*_*Freq*_ and *EUR*_*Freq*_), distance to the *GATM* transcription start site (TSS), and associations with *GATM* expression for lead variants than span the *GATM* locus (Table S2). There were only two other studies that found Scr-associated variants with effect alleles at high *AFR*_*Freq*_ and low *EUR*_*Freq*_, i.e. African ancestry-enriched variants [42, 48]. However, these two variants were much further from the *GATM* TSS, located 25kb and 78kb downstream of the gene, and their associations with *GATM* expression were weaker than seen for the variants discovered here. The lead variants discovered here via standard GWAS (rs2467850) and ancestry-specific GWAS (rs8024550) show the lowest *EUR*_*Freq*_ (≤0.001), are closest to the TSS, located in the proximal promoter (−1365) and 5’ UTR (+61), and show the strongest associations with *GATM* expression (*β*=0.72 and *β*=0.70). Thus, the variants discovered here can be considered to best explain increased Scr levels in African ancestry patients mediated by upregulation of *GATM*.

Our focus on an admixed African-European ancestry cohort from the All of Us Research Program provides several advantages. First, it powers the ancestry-informed association approaches used here, admixture mapping and ancestry-specific GWAS [25, 28], both of which replicated the standard GWAS findings and underscored the link between African ancestry-enriched variants (haplotypes) and increased Scr levels (Figure 2 and Table 1). Second, it serves as an internal control for potential non-genetic confounders, e.g. social determinants of health or environmental exposures, that may also differ between ancestry groups [49, 50]. Indeed, we see no evidence of genomic inflation owing to population structure in the GWAS conducted here. Third, it allows for the evaluation of the effect of ancestry-specific variants and haplotypes on the kind of admixed genetic background seen in African ancestry participants. Finally, the All of Us cohort draws volunteer participants from the same US population that has been at the center of the debate on the use of race in clinical algorithms [9, 10, 34]. As such, we consider the results obtained here to be directly relevant to eGFR equations.

This study is limited by its reliance on the All of Us Research Program, which recruits volunteer participants and is thus not a nationally representative sample. As such, GWAS effect sizes estimated on our study cohort may not be externally valid to the US population [51, 52]. This concern is mitigated by the similar Scr associations seen at the *GATM* locus for previous GWAS [42, 48]. In addition, given that this is an observational study, there could be hidden confounding variables associated with Scr levels that have not be considered here, such as diet, lifestyle, or supplement use. Nevertheless, previous studies have shown that factors like diet, height, weight, and socioeconomic variables are not relevant to Scr associations with genetic ancestry [7, 8, 15]. Finally, the study may be limited by the use of Scr measurements taken from participant EHR, which could lead to differences based on laboratory measurement bias. This challenge is underscored by the fact that some Scr levels were reported as *μ*mol/L while others were reported as mg/dL, necessitating unit conversion prior to analysis. In addition, unhealthy participants may be more likely to have laboratory Scr measures in their EHR; although, this study focused on healthy individuals by excluding participants with kidney disease diagnoses. Another challenge related to EHR was multiple Scr measures for individual patients, for which we took median values after eliminating outliers. Scr levels measured at enrollment using standard procedures, such as done for the UK Biobank, may be more consistent among participants [53].

## Conclusion

The finding of African ancestry-enriched variants associated with Scr in African ancestry participants points to the use of precision medicine, as opposed to race-based medicine, when it comes to diagnostic algorithms in general and eGFR calculation in particular. Allele frequency differences for variants that are associated with Scr will be, at best, imperfectly captured by socially defined racial categories or genome-wide ancestry percentages for that matter. Rather, individual-level genetic information, such as captured by the Scr polygenic score (*PGS*_*SCR*_) generated here, will be far more informative as variables for eGFR equations. We propose that the GWAS summary statistics generated here, and in other Scr association studies, be leveraged to generate polygenic scores that can be used to adjust eGFR equations and predict that this approach will yield more accurate results than the prior race-adjusted or the currently used race-neutral equations.

## Data Availability

This study used data from version 7 of the All of Us Research Program's Controlled Tier Dataset. All the data and analysis code (upon request) are available to authorized users on the Researcher Workbench.

https://workbench.researchallofus.org/

## List of abbreviations

Scr: Serum creatinine
eGFR: Estimated glomerular filtration rate
mGFR: Measured glomerular filtration rate
MDRD: Modification of Diet in Renal Disease
CKD-EPI: Chronic Kidney Disease Epidemiology Collaboration
All of Us: All of Us Research Program
EHR: Electronic health records
GWAS: Genome-wide association study
srWGS: Short-read whole genome sequencing
ACAF: Allele count/allele frequency
LD: Linkage disequilibrium
PC: Principal component
Q-Q: Quantile-quantile
*ß*: Beta
PGS: Polygenic score
TSS: Transcription start site
UTR: Untranslated region
AFR: African
EUR: European
EAS: East Asian
AA: African American
NA: Native American

## Declarations

### Ethics approval and consent to participate

The All of Us IRB has approved of the protocols and materials used by the program before participants were enrolled in the All of Us research program.

### Consent for publication

Not applicable.

### Authors’ contributions

S.S., L.M.R., and I.K.J. conceptualized this study and prepared the manuscript. L.M.R. and I.K.J. provided supervision and funding for the All of Us Researcher Workbench analysis. S.S. and I.K.J. designed the analysis. S.S. and C.A.A. performed all the analyses. All authors read and approved the final manuscript.

## Acknowledgements

We sincerely acknowledge All of Us participants as research partners and their contributions, without whom this research would not have been possible. We are also grateful to the National Institutes of Health’s All of Us Research Program for providing us with the genomic and health data analyzed in this study.

