## Supplementary Information for "African ancestry-enriched variants in the *GATM* gene are associated with elevated serum creatinine levels"

Figure S1. **Analysis flowchart.** Inclusion and exclusion criteria are shown for each step along with the number of participants retained. The final cohort of 18,979 participants was used for GWAS, local ancestry inference, admixture mapping, and ancestry-specific GWAS.

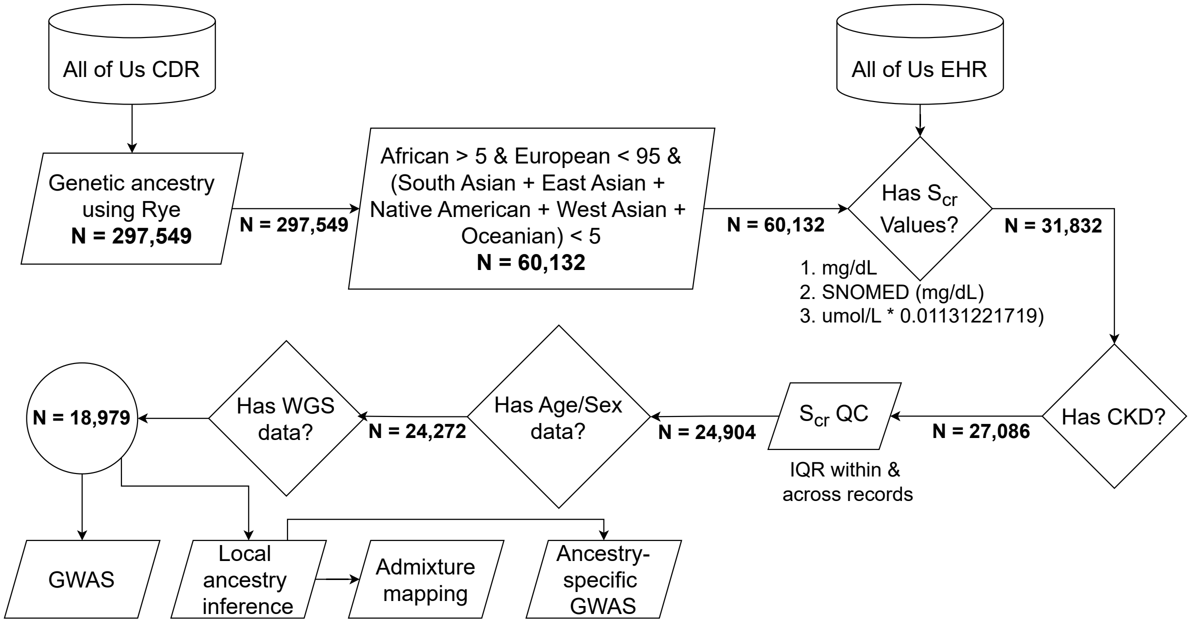

Figure S2. **Serum creatinine levels.** Distribution of serum creatinine (Scr) levels shown through histograms stratified by sex.

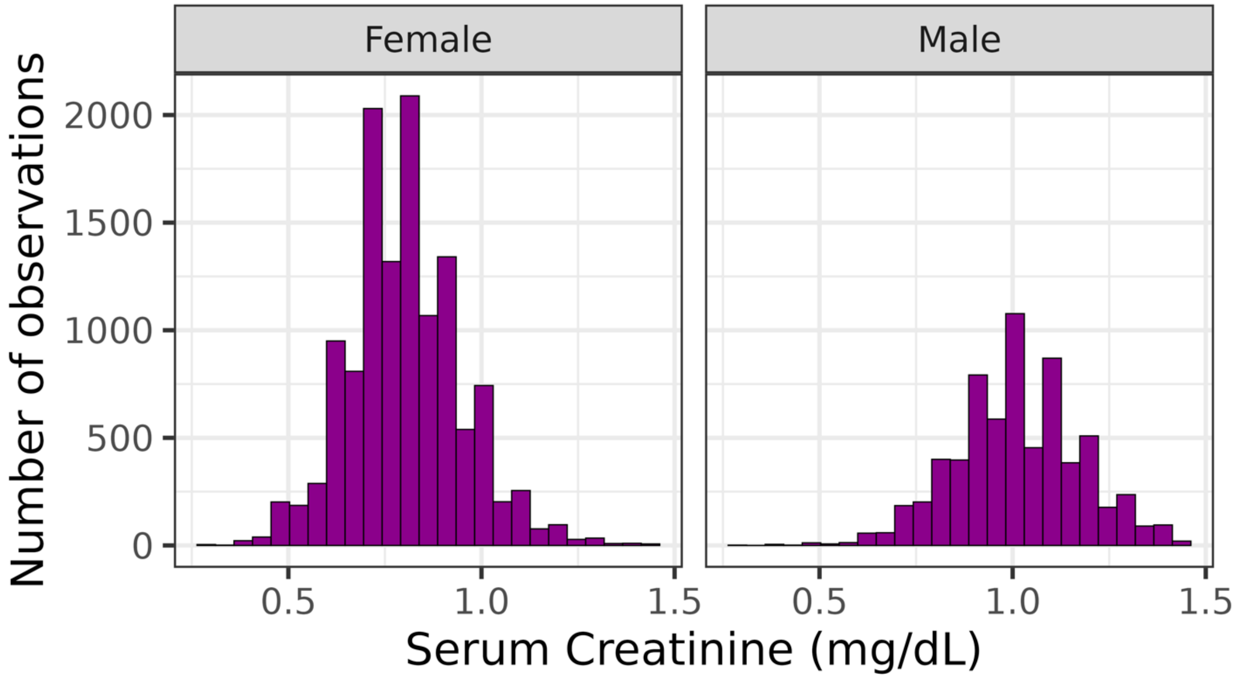

Figure S3. **Q-Q plots.** Quantile-quantile plots comparing the observed p-values vs the expected p-values for (A) standard GWAS, and (B) African ancestry-specific GWAS.

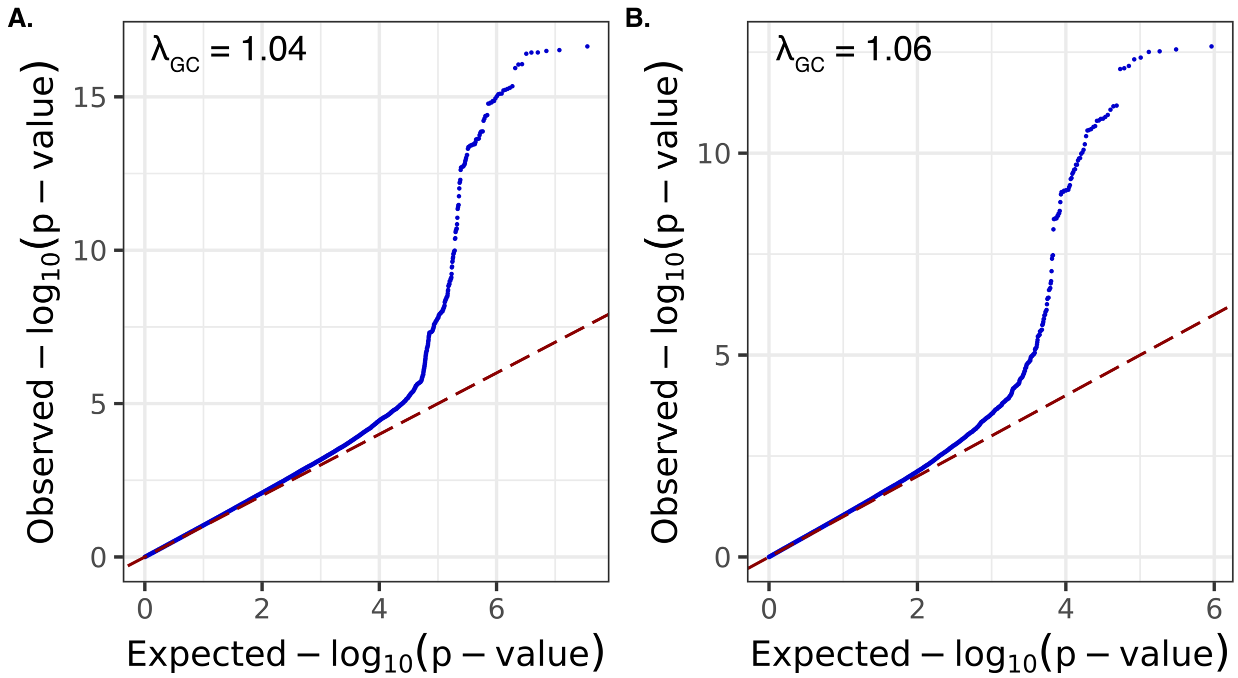

Figure S4.  **Conditional GWAS.** Locus 15q23 (45.3Mb–45.5Mb), significantly associated in the standard GWAS, loses significance only when conditioned on the lead variant rs2467850, but remains significant when conditioned on the second lead variant rs55673230.

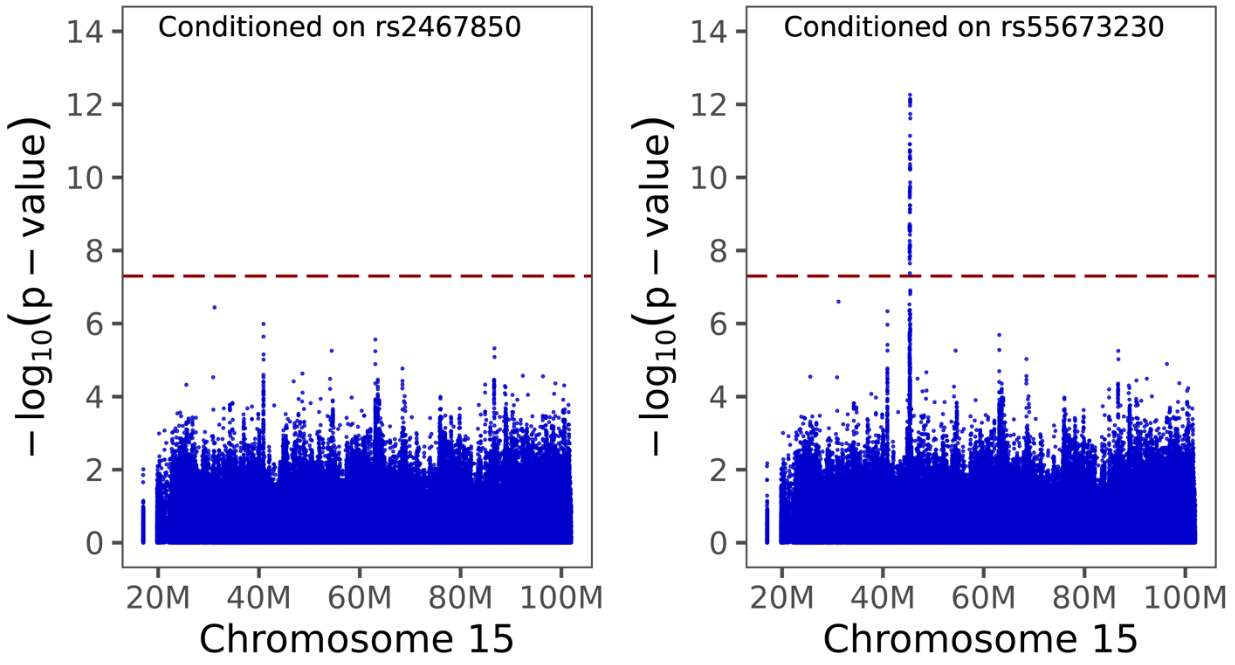

Figure S5. **Conditional admixture mapping.** Haplotypes within and around the *GATM* gene lose significance in the admixture mapping analysis when the model is conditioned on the GWAS lead variant rs2467850.

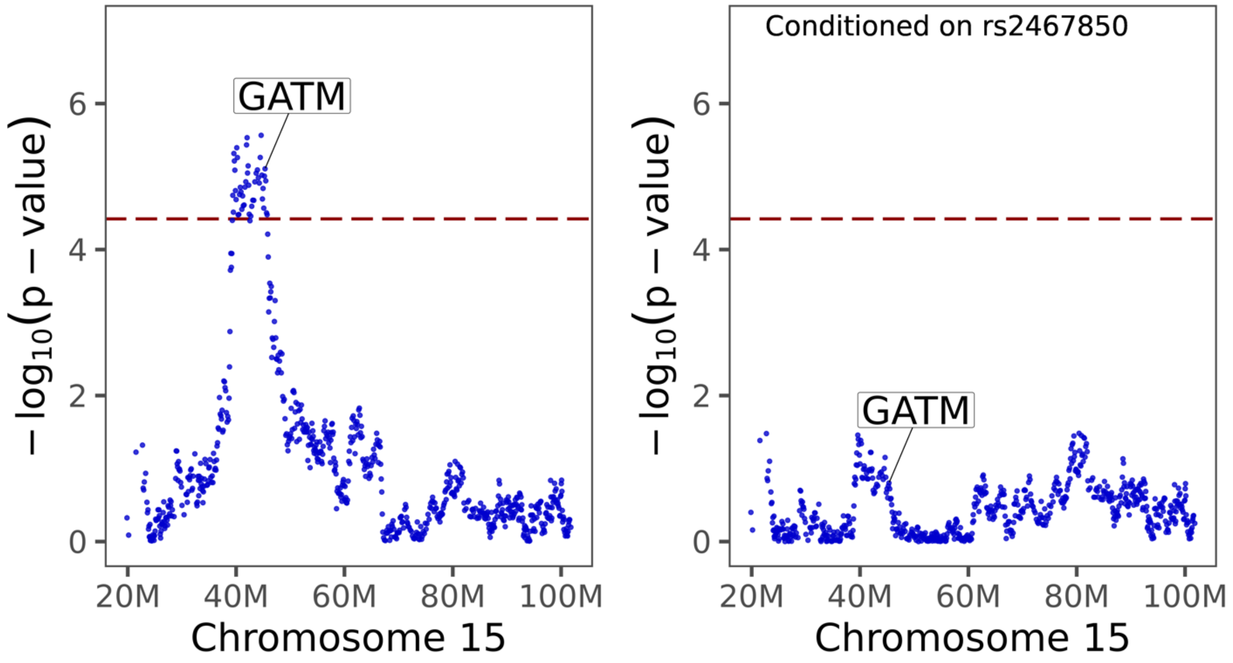

Supplementary Table S1. ***GATM* eQTL in the Scr credible variant set.** African American whole blood eQTL summary statistics for Scr credible variants. TSS dist. is the distance to the *GATM* transcription start site. $AFR_{Freq}$ and $EUR_{Freq}$ are the alternate allele frequencies for the Gnomad African/African American and European (non-Finnish) ancestry groups, respectively.

| ID | CHR | POS | TSS dist. | REF | ALT | BETA | SE | p | $AFR_{Freq}$ | $EUR_{Freq}$ |
|---|---|---|---|---|---|---|---|---|---|---|
| rs1145096 | 15 | 45338576 | 39,968 | A | T | 0.59 | 0.0383 | $7.08\times10^{-46}$ | 0.438 | 0.001 |
| rs1706802 | 15 | 45341646 | 36,898 | G | A | 0.59 | 0.0384 | $1.33\times10^{-46}$ | 0.434 | 0.001 |
| rs1153856 | 15 | 45367576 | 10,968 | T | A | 0.71 | 0.0348 | $1.59\times10^{-73}$ | 0.473 | 0.002 |
| rs8024550 | 15 | 45378483 | 61 | A | C | 0.7 | 0.0369 | $3.79\times10^{-65}$ | 0.424 | 0.001 |
| rs2467850 | 15 | 45379909 | -1,365 | C | T | 0.72 | 0.0371 | $1.25\times10^{-67}$ | 0.413 | 0.001 |
| rs1834686 | 15 | 45402570 | -24,026 | G | T | 0.71 | 0.0367 | $3.81\times10^{-67}$ | 0.422 | 0.001 |
| rs1346265 | 15 | 45403414 | -24,870 | C | T | 0.71 | 0.0351 | $1.06\times10^{-72}$ | 0.472 | 0.001 |
| rs1153848 | 15 | 45403676 | -25,132 | A | G | 0.71 | 0.0369 | $1.05\times10^{-65}$ | 0.414 | 0.001 |
| rs2413765 | 15 | 45406220 | -27,676 | G | A | 0.7 | 0.0369 | $9.44\times10^{-66}$ | 0.413 | 0.001 |
| rs1145075 | 15 | 45407854 | -29,310 | A | G | 0.71 | 0.0369 | $6.32\times10^{-66}$ | 0.415 | 0.001 |
| rs34691692 | 15 | 45412398 | -33,854 | CA | C | 0.72 | 0.0387 | $1.05\times10^{-62}$ | 0.434 | 0.002 |
| rs2467859 | 15 | 45419421 | -40,877 | C | T | 0.7 | 0.0367 | $1.62\times10^{-65}$ | 0.416 | 0.001 |
| rs2453547 | 15 | 45425847 | -47,303 | C | T | 0.7 | 0.0368 | $4.66\times10^{-65}$ | 0.414 | 0.001 |

Supplementary Table S2. **Previously discovered Scr-associated variants at the *GATM* locus.** Lead variants previously found to be associated with Scr or eGFR and spanning the *GATM* locus on chromosome 15 are shown along with the lead variants discovered here using standard and ancestry-specific GWAS (highlighted in yellow). The location of the *GATM* transcript start site (TSS) is shown relative to the variant positions; positive (+) distances are downstream from the TSS, and negative (-) distances are upstream from the TSS. African American whole blood eQTL summary statistics for *GATM* are shown for each variant. TSS dist. is the distance to the *GATM* transcription start site. $AFR_{Freq}$ and $EUR_{Freq}$ are the alternate allele frequencies for the Gnomad African/African American and European (non-Finnish) ancestry groups, respectively. Cohort ancestry abbreviations from the GWAS catalog are: AA-African American, AFR-African Ancestry, AMR-Admixed American or Hispanic, EAS-East Asian, EUR-European, NA-Native American, SAS-South Asian. PMID are the PubMed publication identifiers for the previous GWAS studies.

| ID | CHR | POS | REF | ALT | $AFR_{Freq}$ | $EUR_{Freq}$ | TSS dist. | eQTL BETA | eQTL SE | eQTL p | COHORT ANCESTRY | PMID |
|---|---|---|---|---|---|---|---|---|---|---|---|---|
| rs1464559 | 15 | 45300689 | A | T | 0.528 | 0.002 | +77855 | 0.495 | 0.039 | 5.29E-34 | AA, AFR | 37644460 |
| rs2433603 | 15 | 45354028 | T | C | 0.495 | 0.002 | +24516 | 0.618 | 0.037 | 5.51E-54 | AFR | 33783510 |
| rs2461702 | 15 | 45355229 | A | G | 0.774 | 0.26 | +23315 | -0.184 | 0.052 | 4.38E-04 | AA,AMR,EAS,EUR,NA | 33418499 |
| rs1145093 | 15 | 45357615 | C | A | 0.842 | 0.376 | +20929 | -0.317 | 0.056 | 1.76E-08 | EUR | 33414548 |
| | | | | | | | | | | | EUR,EAS | 31015462 |
| rs62025168 | 15 | 45360080 | G | A | 0.037 | 0.196 | +18464 | 0.293 | 0.120 | 1.52E-02 | EUR,AFR,SAS | 33462484 |
| rs1145085 | 15 | 45365606 | A | G | 0.784 | 0.262 | +12938 | -0.245 | 0.052 | 2.96E-06 | AA,AMR | 38190104 |
| rs1153855 | 15 | 45368560 | C | G | 0.843 | 0.376 | +9984 | -0.317 | 0.056 | 1.76E-08 | EUR,AMR,AA | 37253714 |
| rs2486274 | 15 | 45374030 | G | T | 0.841 | 0.376 | +4514 | -0.314 | 0.055 | 2.15E-08 | EUR | 36357675 |
| rs8024550 | 15 | 45378483 | A | C | 0.424 | 0.001 | +61 | 0.701 | 0.037 | 3.79E-65 | AA | Ancestry-specific GWAS |
| **GATM TSS chr15 45378544** | | | | | | | | | | | | |
| rs2467850 | 15 | 45379909 | C | T | 0.413 | 0.001 | -1365 | 0.721 | 0.037 | 1.25E-67 | AA | Standard GWAS |
| rs2486272 | 15 | 45380055 | T | C | 0.789 | 0.272 | -1511 | -0.252 | 0.052 | 1.35E-06 | EUR,AA | 31451708 |
| | | | | | | | | | | | AA,AMR,EAS,EUR | 39024449 |
| rs1145084 | 15 | 45383242 | G | A | 0.843 | 0.376 | -4698 | -0.321 | 0.056 | 1.13E-08 | EUR | 34272381 |
| | | | | | | | | | | | EAS,SAS,EUR | 38448586 |
| rs1145077 | 15 | 45391597 | G | T | 0.843 | 0.375 | -13053 | -0.317 | 0.056 | 1.76E-08 | EUR,EAS,AA,SAS,AMR | 31152163 |
